# Predictive validity of the simplified Radiographic Assessment of Lung Edema score for the mortality in critically ill COVID-19 patients with the B.1.617.2 (Delta) variant in Vietnam: a single-centre, cross-sectional study

**DOI:** 10.1101/2024.03.20.24304599

**Authors:** Son Ngoc Do, Tuan Quoc Dang, Chinh Quoc Luong, My Ha Nguyen, Dung Thi Pham, Viet Khoi Nguyen, Tan Dang Do, Thai Quoc Nguyen, Vuong Minh Nong, Khoi Hong Vo, Tan Cong Nguyen, Nhung Hong Khuat, Quynh Thi Pham, Dat Tien Hoang, Anh Diep Nguyen, Phuong Minh Nguyen, Duong Dai Cao, Dung Thuy Pham, Dung Tuan Dang, Dat Tuan Nguyen, Vinh Duc Nguyen, Thuan Quang Le, Hung Duc Ngo, Dung Van Nguyen, Thach The Pham, Dung Tien Nguyen, Nguyen Trung Nguyen, Nhung Thi Huynh, Nga Thu Phan, Cuong Duy Nguyen, Thom Thi Vu, Cuong Duy Do, Chi Van Nguyen, Giap Van Vu, Co Xuan Dao

## Abstract

**Background:** Evaluating the prognosis of COVID-19 patients who may be at risk of mortality using the simple chest X-ray (CXR) severity scoring systems provides valuable insights for treatment decisions. This study aimed to assess how well the simplified Radiographic Assessment of Lung Edema (RALE) score could predict the death of critically ill COVID-19 patients in Vietnam.

**Methods:** From July 30 to October 15, 2021, we conducted a cross-sectional study on critically ill COVID-19 adult patients at an intensive care centre in Vietnam. We calculated the areas under the receiver operator characteristic (ROC) curve (AUROC) to determine how well the simplified RALE score could predict hospital mortality. In a frontal CXR, the simplified RALE score assigns a score to each lung, ranging from 0 to 4. The overall severity score is the sum of points from both lungs, with a maximum possible score of 8. We also utilized ROC curve analysis to find the best cut-off value for this score. Finally, we utilized logistic regression to identify the association of simplified RALE score with hospital mortality.

**Results:** Of 105 patients, 40.0% were men, the median age was 61.0 years (Q1-Q3: 52.0-71.0), and 79.0% of patients died in the hospital. Most patients exhibited bilateral lung opacities on their admission CXRs (99.0%; 100/102), with the highest occurrence of opacity distribution spanning three (18.3%; 19/104) to four quadrants of the lungs (74.0%; 77/104) and a high median simplified RALE score of 8.0 (Q1-Q3: 6.0-8.0). The simplified RALE score (AUROC: 0.747 [95% CI: 0.617-0.877]; cut-off value ≥5.5; sensitivity: 93.9%; specificity: 45.5%; P_AUROC_ <0.001) demonstrated a good discriminatory ability in predicting hospital mortality. After adjusting for confounding factors such as age, gender, Charlson Comorbidity Index, serum interleukin-6 level upon admission, and admission severity scoring systems, the simplified RALE score of ≥5.5 (adjusted OR: 18.437; 95% CI: 3.215-105.741; p =0.001) was independently associated with an increased risk of hospital mortality.

**Conclusions:** This study focused on a highly selected cohort of critically ill COVID-19 patients with a high simplified RALE score and a high mortality rate. Beyond its good discriminatory ability in predicting hospital mortality, the simplified RALE score also emerged as an independent predictor of hospital mortality.

## INTRODUCTION

Since the first reports of coronavirus disease 2019 (COVID-19) cases from Wuhan, a city in the Hubei Province of China, at the end of 2019, cases have emerged across all continents.(1–3) As of February 4, 2024, there have been more than 770 million confirmed cases of COVID-19 worldwide.(4) However, the reported case counts underestimate the overall burden of COVID-19, as only a fraction of acute infections are diagnosed and reported. During the COVID-19 pandemic, most countries lack sufficient diagnostic tools due to the rising daily number of reported cases. Therefore, it becomes crucial to correctly categorize COVID-19 patients based on the severity of their symptoms for efficient resource distribution.(5) Specifically, the value of oxygen saturation in the peripheral blood (SpO_2_) is one of the first measures checked for each patient on admission; it often reflects the degree of lung function impairment. The requirement to transfer patients with COVID-19 to an intensive care unit (ICU) depends mainly on their SpO_2_ and concurrent comorbidities.(6) Some experts have suggested that imaging can help determine this need.

A chest X-ray (CXR) has been suggested as a tool to predict the severity of COVID-19 by assessing lung involvement and offering insights into the prognosis of the COVID-19 infection. A previous study introduced the CXR scoring system for quantifying the severity and progression of lung abnormalities in COVID-19 pneumonia.(7) This system evaluated the extent of COVID-19 pulmonary abnormalities on CXR using a semiquantitative severity score ranging from 0 to 3, with 1-point increments, across six lung zones (total range 0–18), correlated these findings with clinical data, and assessed interobserver agreement. Interestingly, the study revealed a moderate to almost perfect interobserver agreement and identified significant but weak correlations with clinical parameters.(7) As a result, integration of CXR into the classification of COVID-19 patients is a potential avenue for further exploration. Another previous study introduced a CXR scoring system called the Brixia score.(8) This score is a valuable tool for assessing the severity of COVID-19 infection. By evaluating the extent and characteristics of lung abnormalities, the Brixia score offers insights into pulmonary involvement using an 18-point severity scale.(8) While CXR is considered not sensitive in detecting early-stage pulmonary involvement, the Brixia score remains a useful diagnostic tool for identifying the rapid progression of lung abnormalities in COVID-19 patients, particularly in ICUs.(8)

To further understand the radiographic features of COVID-19, a retrospective study examines the progression and severity of CXR findings associated with the disease.(9) Additionally, this study adapts and simplifies the original Radiographic Assessment of Lung Edema (original RALE)(10) score to quantify the extent of infection and calculate a severity score for COVID-19. While 20% of patients did not exhibit any abnormalities on their CXR throughout the illness, this study reveals that common abnormal CXR findings included consolidation and ground-glass opacities.(9) These abnormalities were distributed bilaterally, peripherally, and in the lower lung zones. The CXR severity score progressively intensified during the illness, reaching its peak severity between 10 to 12 days after the onset of symptoms.(9) Although this study reveals that baseline CXR sensitivity is lower than those of real-time reverse transcription-polymerase chain reaction (RT-PCR) tests and computed tomography (CT) scans, it still suggests that CXR have a role in the initial screening for COVID-19 and identifying the rapid progression of lung abnormalities in COVID-19 patients.(9)

Two recent published studies also show that the adaptation and simplification of the original RALE score proved valuable in predicting the mortality of critically ill patients with COVID-19.(11, 12) However, limited data on this simplified score’s external validity are available. Therefore, the present study aimed to assess how well the simplified RALE score could predict the death of critically ill patients with COVID-19 in Vietnam.

## METHODS

### Study design and setting

From July 30 to October 15, 2021, we conducted a cross-sectional study on critically ill COVID-19 patients with the Delta variant at the Intensive Care Centre for the Treatment of Critically Ill Patients with COVID-19 (study centre) in Ho Chi Minh City (HCMC), Vietnam. This centre was a field hospital with 360 beds and was affiliated with the Bach Mai Hospital (BMH) in Hanoi, Vietnam. BMH is designated as the central hospital in northern Vietnam by the Ministry of Health (MOH) of Vietnam.(13) It is a large general hospital with 3,200 beds.

In July 2021, HCMC faced a severe Delta variant COVID-19 outbreak, which led to a high demand for ICUs for critically ill patients.(14, 15) In response, the MOH established four specialized Intensive Care Centres for the Treatment of Critically Ill Patients with COVID-19 on July 30, 2021, which could only receive patients transferred from lower-level hospitals in HCMC. The study centre was under the management of BMH and was called Field Hospital Number 16. The study centre had a capacity of 500 beds at its peak and later reduced it to 360. The study centre stopped operating on April 30, 2022, after the COVID-19 cases declined. The Director of BMH also served as the Director of the study centre. Likewise, two Deputy Directors of the study centre were from the Centre for Emergency Medicine of BMH in Hanoi and the Director Board of Hung Vuong Hospital in HCMC. The study centre had 968 medical staff and volunteers in total, and more than half of them (542 medical staff) were from BMH. They came from different departments and centres of BMH, such as Emergency, Intensive Care, Tropical Diseases, Respiratory, Clinical Nutrition, Nephro-Urology and Dialysis, Neurology, Cardiovascular, Infection Control, Haematology and Blood Transfusion, Radiology, Biochemistry, and Microbiology.

### Participants

This study included patients aged 18 years or older who were critically ill with COVID-19 and presented to the study centre. We defined a case of COVID-19 as a person with laboratory confirmation of COVID-19 infection, regardless of clinical or epidemiological criteria.(16, 17) Laboratory confirmation of COVID-19 infection was based on a positive RT-PCR test using samples obtained from the respiratory tract by nasopharyngeal swab, throat or mouth swab, or tracheal lavage fluid.(18) We excluded patients who did not have CXRs between hospital admission and the end of the first day.

### Data collection

The data for each study patient were recorded from the same unified case record forms (CRFs). Data was entered into the study database by the online password-protected CRF. Patient identifiers were not entered in the database to protect patients’ confidentiality.

### Variables

The CRF contained four sections, which included variables mainly based on the COVID-19 Treatment Guidelines of the National Institutes of Health(19) and collected by fully trained clinicians, such as information on:

(i) The first section focused on baseline characteristics, such as: prehospital care (e.g., prior hospitalization, prehospital airway, prehospital oxygen), demographics (i.e., age and gender), documented comorbidities (e.g., cerebrovascular disease, chronic cardiac failure, coronary artery disease/myocardial infarction, hypertension, chronic obstructive pulmonary disease/asthma, other chronic pulmonary disease, tuberculosis, chronic renal failure, and diabetes mellitus), and details of admission. We also used the 19 comorbidity categories to compute the Charlson Comorbidity Index (CCI) Score, which measures the predicted mortality rate based on the presence of comorbidities.(20) A score of zero indicates that no comorbidities were detected; the higher the score, the higher the expected mortality rate is.(20–22)

(ii) The second section comprised characteristics upon admission, such as vital signs (e.g., heart rate, respiration rate, arterial blood pressure, and body temperature), laboratory parameters (e.g., interleukin-6 (IL-6)), chest X-ray (CXR) findings (e.g., bilateral opacities, number of involved quadrants, and severity of COVID-19 based on lung involvement), gas exchange (e.g., the partial pressure of oxygen (PaO_2_) and carbon dioxide (PaCO_2_) in the arterial blood, the acidity of the blood (pH), and the ratio of partial pressure of oxygen in arterial blood to the fraction of inspiratory oxygen concentration (PaO_2_/FiO_2_)), and severity scoring systems (e.g., Sequential Organ Failure Assessment (SOFA) score(23), Acute Physiology and Chronic Health Evaluation II (APACHE II) score(24), and Confusion, Urea >7 mmol/L, Respiratory Rate ≥30 breaths/min, Blood Pressure <90 mm Hg (Systolic) or <60 mm Hg (Diastolic), Age ≥65 Years (CURB-65) score(25)). Serum IL-6 was measured using the Elecsys IL-6 immunoassay (Roche^®^ Cobas E411 analyser), an in vitro diagnostic test for quantitatively determining IL-6 in human serum and plasma. The measuring range is from 1.5 – 5000pg/mL. Data on the IL-6 levels were collected prospectively on a CRF from hospital admission until the end of the first day. In the present study, we defined the IL-6 level as 5000pg/mL when values were above the measuring range (>5000pg/mL). Prospectively, we collected all necessary data elements for calculating the SOFA score, APACHE II score, and CURB-65 score within the time window from admission until 24 hours later, entered these data into an online CRF, and stored them in a database for the subsequent analysis. The CXRs were performed for severely and critically ill patients with COVID-19 in a single frontal projection with an anteroposterior view by using a compact and portable X-ray device (FDR nano, Model/Serial: DR-XD 1000, Fujifilm Techno Products Co., Ltd) conveniently brought to the patient’s bedside in the hospital or emergency room. The X-ray tube was connected to a flexible arm, allowing the technician to position it over the patient and place an X-ray film holder or image recording plate underneath. In hospitalized patients with COVID-19, we performed a portable CRX as the initial evaluation for pulmonary complications and lung involvement. Chest computed tomography (CT) scans were reserved for specific situations that might change clinical management, partly to minimize infection control issues related to transport. These practices align with the recommendations from the American College of Radiology(26). The evaluation of CXR findings, which were taken from hospital admission until the end of the first day and included bilateral opacities, the extent of lung involvement across quadrants, and the severity of COVID-19 based on lung assessments, was a two-step process. Initially, a radiology resident in their third year reviewed the CXRs. Following this, a thoracic radiologist with ten years of experience independently validated the findings. Both professionals were unaware of other clinical variables and outcomes. In case of any discrepancies identified in the images, they collaborated to reach a consensus. We adapted and simplified the original RALE score(10) to create a simplified RALE score for assessing the severity of COVID-19 based on lung involvement. This approach has also been used in prior published studies to quantify the extent of pulmonary infection in COVID-19 cases.(9, 11, 12, 17) In a frontal CXR, the simplified RALE scoring system assigns a score to each lung, ranging from 0 to 4. This score reflects the attendance of specific abnormalities. The system divides each lung into four zones, and abnormalities such as consolidation, ground-glass opacification, and reticular interstitial thickening are assigned a score of 1 in each zone (S1 Fig in S1 File). A score of 0 indicates no involvement, while a score of 1 corresponds to less than 25% involvement, a score of 2 represents 25% to less than 50% involvement, a score of 3 signifies 50% to less than 75% involvement, and a score of 4 indicates 75% or greater involvement. The overall severity score is the sum of points from both lungs, with a maximum possible score of 8. Based on the simplified RALE score upon admission, the patients were classified into three severity groups as follows: mild (a score of 1 to 2), moderate (a score of 3 to 6), and severe (a score of 7 to 8).

(iii) The third section captured life-sustaining treatments provided during the ICU stay, such as respiratory support on the first day of admission (e.g., oxygen supplements and mechanical ventilation) and adjunctive therapies (e.g., prone positioning, recruitment maneuvers, extracorporeal membrane oxygenation, antiviral drugs, antibiotics, corticosteroids, heparin, antiplatelet drugs, novel oral anticoagulants, recombinant humanized anti-IL-6 receptor monoclonal antibody, continuous sedation, continuous neuromuscular blocking agents, renal replacement therapy, cytokine adsorption therapy, tracheostomy, and inhaled vasodilators).

(iv) The fourth section is concerned with complications (e.g., hospital-acquired pneumonia (HAP), hypoxemic respiratory failure/ARDS, diffuse alveolar damage, secondary bacterial infections, sepsis and septic shock, cardiac injury, arrhythmia, acute kidney injury, liver dysfunction, multi-organ failure, thromboembolism, gastrointestinal bleeding, and pneumothorax/pneumomediastinum) and clinical outcomes (e.g., hospital mortality). HAP is defined as pneumonia that occurs 48 hours or more after admission and does not appear to be incubating at the time of admission.(27) To diagnose and classify ARDS, we applied the Berlin criteria, which categorize ARDS severity based on the PaO_2_/FiO_2_ ratio as follows: mild (200 < PaO_2_/FiO_2_ ≤ 300 mmHg), moderate (100 < PaO_2_/FiO_2_ ≤ 200 mmHg), and severe (PaO_2_/FiO_2_ ≤ 100 mmHg), with a minimum positive end-expiratory pressure (PEEP) of 5 cmH_2_O applied to the lungs at the end of each breath.(28, 29). Additionally, septic shock is identified as a clinical construct of sepsis characterized by persisting hypotension requiring vasopressors to maintain a mean arterial pressure ≥ 65 mmHg, along with a serum lactate level > 2 mmol/L (18 mg/dL) despite adequate volume resuscitation.(30) Lastly, thromboembolism encompasses venous thromboembolism (i.e., deep vein thrombosis and pulmonary embolism), arterial events (i.e., stroke, limb ischemia, and myocardial infarction), and microvascular thrombosis (e.g., microvascular thrombosis in the lungs).(19) We followed all patients till hospital discharge or death in the ICU/hospital, whichever was earliest.

### Outcome measures

The primary outcome was hospital mortality, which we defined as death from any cause during the hospitalization. We also examined the secondary outcomes, such as complications (e.g., ARDS) and hospital lengths of stay.

### Sample size

In this cross-sectional study, the primary outcome was hospital mortality. Therefore, we used the formula to find the minimum sample size for estimating a population proportion with a confidence level of 90%, a confidence interval (margin of error) of ±8.02%, and an assumed population proportion of 54.64%, based on the hospital mortality (54.64%) reported in a previously published study(31). As a result, our sample size should be at least 105 patients, which might be large enough to reflect a normal distribution.

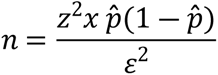

where:

*z is the z score* (*z score for a* 90% *confidence level is* 1.65)

*ε is the margin of error* (*ε for a confidence interval of* ± 8.02% *is* 0.0802)

*p̂ is the population proportion* (*p̂ for a population proportion of* 54.64% *is* 0.5464)

*n is the sample size*

### Statistical analyses

We used IBM^®^ SPSS^®^ Statistics 22.0 (IBM Corp., Armonk, United States of America) and Analyse-it statistical software (Analyse-it Software, Ltd., Leeds, United Kingdom) for data analysis. We report the data as numbers (no.) and percentages (%) for categorical variables and medians and interquartile ranges (Q1-Q3) or means and standard deviations (SDs) for continuous variables. Furthermore, comparisons were made between survival and death in the hospital for each variable using the Chi-squared or Fisher’s exact test for categorical variables and the Mann–Whitney U test, Kruskal–Wallis test, or one-way analysis of variance for continuous variables.

To evaluate how well the simplified RALE score, CCI score, admission serum IL-6 level, SOFA score, APACHE II score, and CURB-65 score could differentiate between patients who survived and those who died in the hospital, we plotted receiver operator characteristic (ROC) curves and calculated areas under the ROC curve (AUROCs) for each score or level. We also used the ROC curve analysis to determine the best cut-off value for each score or level, which was the point that gave the highest Youden’s index (i.e., sensitivity + specificity - 1). Then, we divided the patients into two groups based on their score or level: one that was less than the cut-off or another greater than or equal to the cut-off value. Additionally, we compared the AUROCs of simplified RALE score with those of the serum IL-6 level, CCI score, SOFA score, APACHE II score, and CURB-65 score to see which one was better at predicting deaths in the hospital using the Z-statistics. Finally, we calculated correlation coefficients (Rs) using Spearman’s rho to explore the relationship between simplified RALE score and each score or level.

We assessed the association of simplified RALE Score with hospital mortality using logistic regression analysis. To reduce the number of predictors and the multicollinearity issue and resolve the overfitting, we used different methods to select variables as follows: (a) we put all variables of demographics, documented comorbidities, clinical characteristics, CXR findings, laboratory investigations, gas exchange, severity scoring systems, first-day respiratory support, adjunctive therapies, complications into the univariable logistic regression model; (b) we selected variables if the P-value was <0.05 in the univariable analysis between death and survival in the hospital, as well as those that are clinically crucial, to put in the multivariable logistic regression model. These variables included demographics (i.e., age ≥ 60 years, gender (male)), documented comorbidities (i.e., CCI score ≥cut-off value), CXR findings (i.e., simplified RALE Score ≥cut-off value), laboratory investigations (i.e., admission serum IL-6 level ≥cut-off value), severity scoring systems (i.e., SOFA score ≥ cut-off value, CURB-65 score ≥cut-off value). Using a stepwise backward elimination method, we started with the full multivariable logistic regression model that included the selected variables. This method then deleted the least statistically significant variables stepwise from the full model until all remaining variables were independently associated with hospital mortality in the final model. We presented the odds ratios (ORs) and 95% confidence intervals (CIs) in the univariable logistic regression model and the adjusted ORs (AORs) and 95% CIs in the multivariable logistic regression model.

The significance levels were two-tailed for all analyses, and we considered the P <0.05 as a statistically significant value.

### Ethical issues

This study was approved by the Scientific and Ethics Committees of Bach Mai Hospital (Approval number: 3412/QĐ-BM) and conducted according to the principles of the Declaration of Helsinki. The Bach Mai Hospital Scientific and Ethics Committees waived the written informed consent for this non-interventional study. Verbal informed consents were directly obtained from patients or, when unavailable, from family members over the phone or at the Intensive Care Centre for the Treatment of Critically Ill Patients with COVID-19 and witnessed by the on-duty medical staff. Public notification of this study was made by published posting, according to the Strengthening the Reporting of Observational Studies in Epidemiology (STROBE): Explanation and Elaboration - the STROBE Statement - Checklist of items that should be included in reports of cross-sectional studies. The authors who performed the data analysis kept the data set in password-protected systems and only presented anonymized data.

## RESULTS

### Baseline characteristics and clinical outcomes

Of 105 patients, 40.0% were men, and the median age was 61.0 years (Q1-Q3: 52.0-71.0) (Table 1). Upon admission, nearly all patients (99.0%; 100/102) exhibited bilateral lung opacities on their CXR (Table 2). These opacities were most frequently distributed across three (18.3%; 19/104) and four quadrants of lungs (74.0%; 77/104). Additionally, the patients had a high median simplified RALE score of 8.0 (Q1-Q3: 6.0-8.0) (Table 2) and a low mean PaO_2_/FiO_2_ ratio of 101.66 (SD: 58.20) (Table 2). Elevated mean serum IL-6 level was observed upon admission (99.76pg/mL; SD: 174.33) (Table 2). The median SOFA score was 4.0 (Q1-Q3: 2.0-5.0), median APACHE II score was 8.0 (Q1-Q3: 3.0-12.0), and median CURB-65 score was 1.0 (Q1-Q3: 1.0-2.0) within the time window from admission until 24 hours later (Table 2). In general, 79.0% (83/105) of the patients died in the hospital (Tables 1 and 2). As shown in Tables 1 and 2 and S1 to S5 Tables (S1 File), we also compared the factors between survivors and non-survivors in the hospital, such as prehospital care, demographics, comorbidities, vital signs, laboratory investigations, CXR findings, severity scoring systems, treatments, and complications.

**Table 1.** Baseline characteristics of critically ill COVID-19 patients with the Delta variant, according to hospital survivability.

| Variables | All cases<br>n=105 | Survived<br>n=22 | Died<br>n=83 | P value<sup>a</sup> |
| --- | --- | --- | --- | --- |
| **Demographics** |  |  |  |  |
| Age (year), median (Q1-Q3) | 61.0 (52.0-71.0) | 53.5 (48.5-62.0) | 62.0 (54.0-73.0) | 0.009 |
| Gender (male), no. (%) | 42 (40.0) | 6 (27.3) | 36 (43.4) | 0.171 |
| **Comorbidities** |  |  |  |  |
| Cerebrovascular disease, no. (%), n=78 | 5 (6.4) | 0 (0.0) | 5 (7.7) | 0.583 |
| Chronic cardiac failure, no. (%), n=78 | 4 (5.1) | 2 (15.4) | 2 (3.1) | 0.127 |
| Coronary artery disease/MI, no. (%), n=78 | 4 (5.1) | 1 (7.7) | 3 (4.6) | 0.525 |
| Hypertension, no. (%), n=78 | 50 (64.1) | 7 (53.8) | 43 (66.2) | 0.528 |
| COPD/asthma, no. (%), n=78 | 1 (1.3) | 0 (0.0) | 1 (1.5) | >0.999 |
| Other chronic pulmonary disease, no. (%), n=78 | 1 (1.3) | 0 (0.0) | 1 (1.5) | >0.999 |
| Tuberculosis, no. (%), n=78 | 1 (1.3) | 0 (0.0) | 1 (1.5) | >0.999 |
| Chronic renal failure, no. (%), n=78 | 1 (1.3) | 0 (0.0) | 1 (1.5) | >0.999 |
| Diabetes mellitus, no. (%), n=78 | 40 (51.3) | 5 (38.5) | 35 (53.8) | 0.311 |
| CCI score, median (Q1-Q3), n=102 | 2.0 (1.0-4.0) | 1.0 (0.0-2.5) | 2.0 (1.0-4.0) | 0.023 |
| **Vital signs upon admission** |  |  |  |  |
| HR (beats/min), median (Q1-Q3), n=99 | 92.0 (80.0-100.0) | 90.0 (84.75-100.75) | 95.0 (80.0-100.0) | 0.871 |
| RR (breaths/min), median (Q1-Q3), n=97 | 26.0 (25.0-30.0) | 25.0 (22.0-27.0) | 28.0 (25.0-31.5) | 0.006 |
| Systolic BP (mmHg), mean (SD), n=99 | 124.97 (17.58) | 127.59 (15.03) | 124.22 (18.27) | 0.573 |
| Diastolic BP (mmHg), mean (SD), n=98 | 75.91 (11.51) | 79.0 (8.40) | 75.06 (12.13) | 0.195 |
| Body temperature (°C), mean (SD), n=95 | 36.95 (0.37) | 36.94 (0.15) | 36.95 (0.41) | 0.390 |

<sup>a</sup> The comparison between patients who survived and those who died in the hospital.
**Abbreviations:** **BP**, blood pressure; **CCI**, Charlson Comorbidity Index; **COPD**, chronic obstructive pulmonary disease; **HR**, heart rate; **MI**, myocardial infarction; **no.**, number of patients; **Q**, quartile; **RR**, respiration rate; **SD**, standard deviation.

**Table 2.**
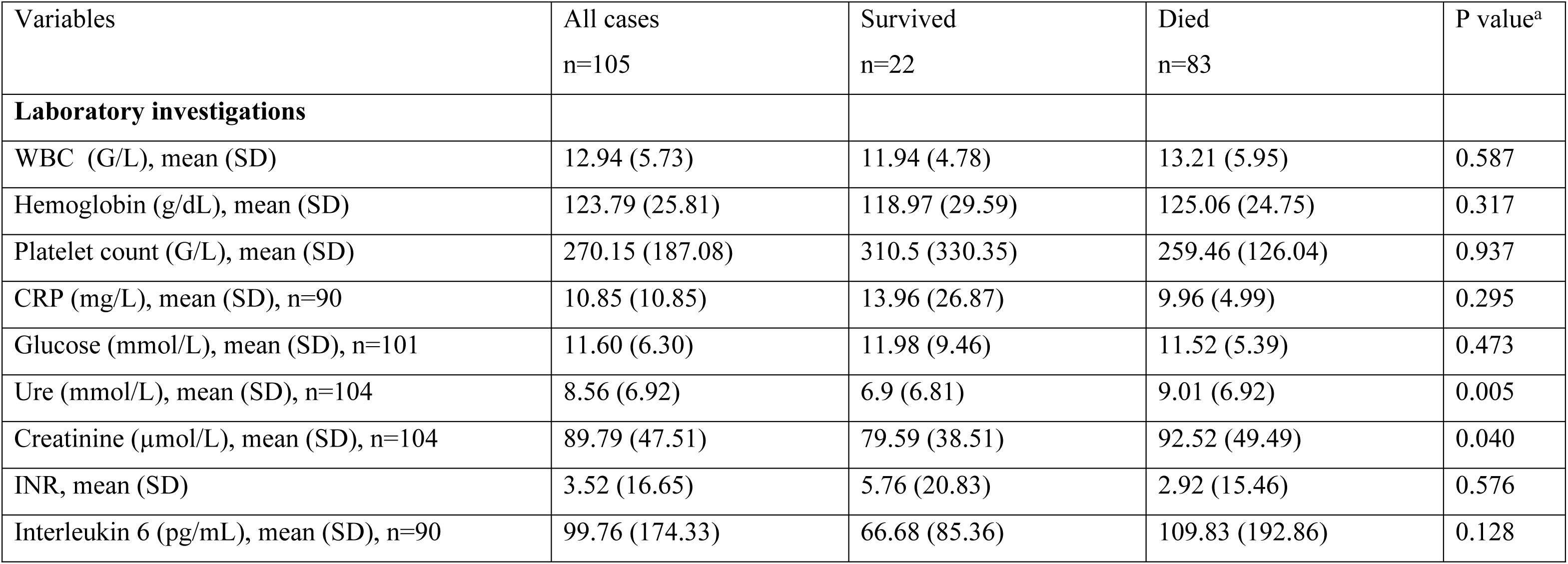
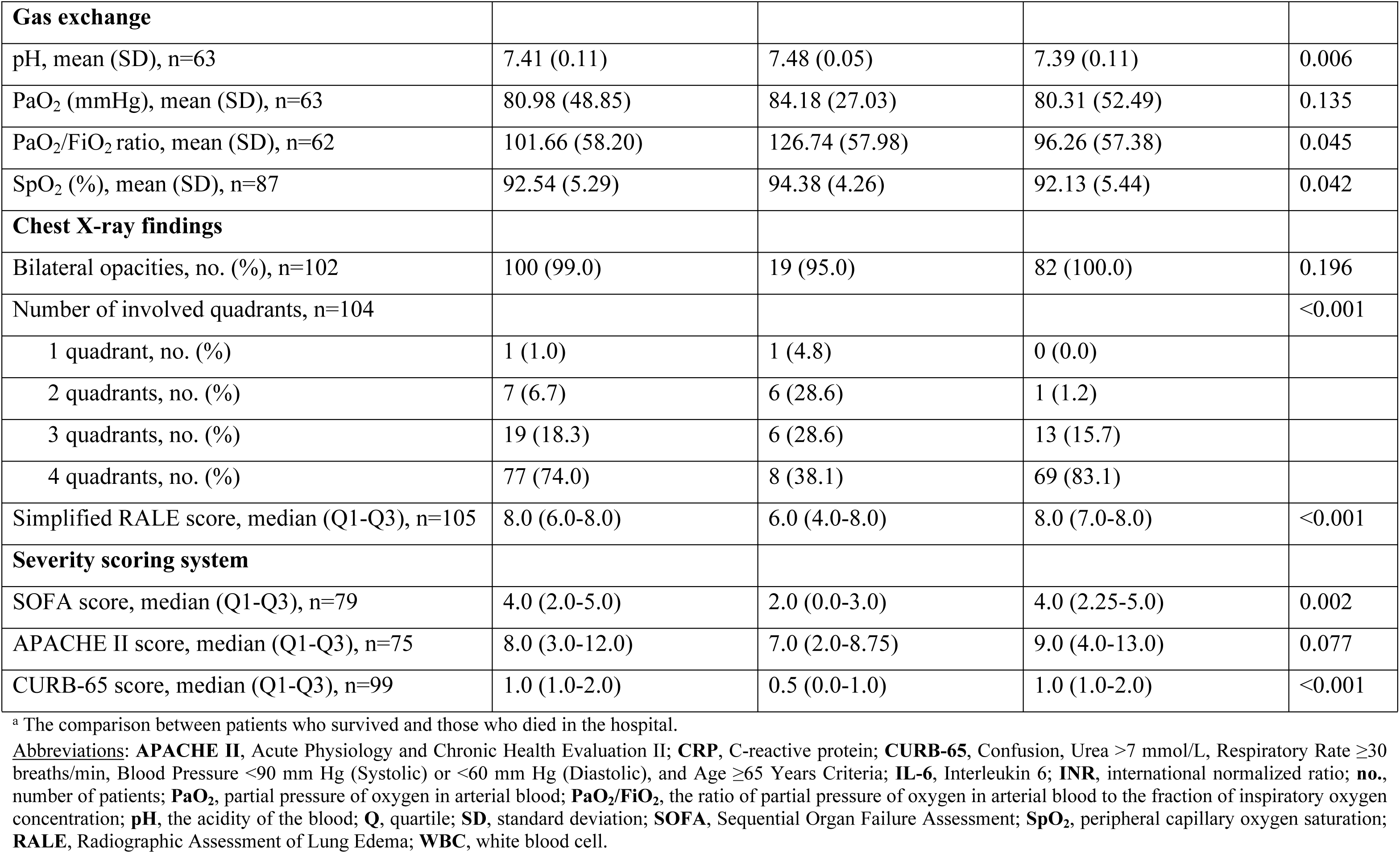
Laboratory investigations, gas exchange, chest X-ray findings, and severity scoring systems of critically ill COVID-19 patients with the Delta variant upon admission, according to hospital survivability.

| Variables | All cases<br>n=105 | Survived<br>n=22 | Died<br>n=83 | P value<sup>a</sup> |
| --- | --- | --- | --- | --- |
| **Laboratory investigations** |  |  |  |  |
| WBC (G/L), mean (SD) | 12.94 (5.73) | 11.94 (4.78) | 13.21 (5.95) | 0.587 |
| Hemoglobin (g/dL), mean (SD) | 123.79 (25.81) | 118.97 (29.59) | 125.06 (24.75) | 0.317 |
| Platelet count (G/L), mean (SD) | 270.15 (187.08) | 310.5 (330.35) | 259.46 (126.04) | 0.937 |
| CRP (mg/L), mean (SD), n=90 | 10.85 (10.85) | 13.96 (26.87) | 9.96 (4.99) | 0.295 |
| Glucose (mmol/L), mean (SD), n=101 | 11.60 (6.30) | 11.98 (9.46) | 11.52 (5.39) | 0.473 |
| Ure (mmol/L), mean (SD), n=104 | 8.56 (6.92) | 6.9 (6.81) | 9.01 (6.92) | 0.005 |
| Creatinine (μmol/L), mean (SD), n=104 | 89.79 (47.51) | 79.59 (38.51) | 92.52 (49.49) | 0.040 |
| INR, mean (SD) | 3.52 (16.65) | 5.76 (20.83) | 2.92 (15.46) | 0.576 |
| Interleukin 6 (pg/mL), mean (SD), n=90 | 99.76 (174.33) | 66.68 (85.36) | 109.83 (192.86) | 0.128 |
| **Gas exchange** |  |  |  |  |
| pH, mean (SD), n=63 | 7.41 (0.11) | 7.48 (0.05) | 7.39 (0.11) | 0.006 |
| PaO<sub>2</sub> (mmHg), mean (SD), n=63 | 80.98 (48.85) | 84.18 (27.03) | 80.31 (52.49) | 0.135 |
| PaO<sub>2</sub>/FiO<sub>2</sub> ratio, mean (SD), n=62 | 101.66 (58.20) | 126.74 (57.98) | 96.26 (57.38) | 0.045 |
| SpO<sub>2</sub> (%), mean (SD), n=87 | 92.54 (5.29) | 94.38 (4.26) | 92.13 (5.44) | 0.042 |
| **Chest X-ray findings** |  |  |  |  |
| Bilateral opacities, no. (%), n=102 | 100 (99.0) | 19 (95.0) | 82 (100.0) | 0.196 |
| Number of involved quadrants, n=104 |  |  |  | <0.001 |
| 1 quadrant, no. (%) | 1 (1.0) | 1 (4.8) | 0 (0.0) |  |
| 2 quadrants, no. (%) | 7 (6.7) | 6 (28.6) | 1 (1.2) |  |
| 3 quadrants, no. (%) | 19 (18.3) | 6 (28.6) | 13 (15.7) |  |
| 4 quadrants, no. (%) | 77 (74.0) | 8 (38.1) | 69 (83.1) |  |
| Simplified RALE score, median (Q1-Q3), n=105 | 8.0 (6.0-8.0) | 6.0 (4.0-8.0) | 8.0 (7.0-8.0) | <0.001 |
| **Severity scoring system** |  |  |  |  |
| SOFA score, median (Q1-Q3), n=79 | 4.0 (2.0-5.0) | 2.0 (0.0-3.0) | 4.0 (2.25-5.0) | 0.002 |
| APACHE II score, median (Q1-Q3), n=75 | 8.0 (3.0-12.0) | 7.0 (2.0-8.75) | 9.0 (4.0-13.0) | 0.077 |
| CURB-65 score, median (Q1-Q3), n=99 | 1.0 (1.0-2.0) | 0.5 (0.0-1.0) | 1.0 (1.0-2.0) | <0.001 |

<sup>a</sup> The comparison between patients who survived and those who died in the hospital.
**Abbreviations:** **APACHE II**, Acute Physiology and Chronic Health Evaluation II; **CRP**, C-reactive protein; **CURB-65**, Confusion, Urea >7 mmol/L, Respiratory Rate ≥30 breaths/min, Blood Pressure <90 mm Hg (Systolic) or <60 mm Hg (Diastolic), and Age ≥65 Years Criteria; **IL-6**, Interleukin 6; **INR**, international normalized ratio; **no.**, number of patients; **PaO<sub>2</sub>**, partial pressure of oxygen in arterial blood; **PaO<sub>2</sub>/FiO<sub>2</sub>**, the ratio of partial pressure of oxygen in arterial blood to the fraction of inspiratory oxygen concentration; **pH**, the acidity of the blood; **Q**, quartile; **SD**, standard deviation; **SOFA**, Sequential Organ Failure Assessment; **SpO<sub>2</sub>**, peripheral capillary oxygen saturation; **RALE**, Radiographic Assessment of Lung Edema; **WBC**, white blood cell.

### Overall predictive performance of simplified RALE score and other severity scoring systems

Unlike the PaO_2_/FiO_2_ ratio (AUROC: 0.306 [95% CI: 0.124-0.487]; cut-off value ≥46.5%; sensitivity: 98.0%; specificity: 9.1%; P_AUROC_ =0.045), serum IL-6 level (AUROC: 0.610 [95% CI: 0.459-0.761]; cut-off value ≥15.8pg/mL; sensitivity: 84.1%; specificity: 42.9%; P_AUROC_ =0.128), and APACHE II score (AUROC: 0.645 [95% CI: 0.504-0.785]; cut-off value ≥11.5; sensitivity: 33.9%; specificity: 93.8; P_AUROC_ =0.078) (S6 Table in S1 File), the simplified RALE score (AUROC: 0.747 [95% CI: 0.617-0.877]; cut-off value ≥5.5; sensitivity: 93.9%; specificity: 45.5%; P_AUROC_ <0.001) demonstrated the good discriminatory ability in predicting hospital mortality (Fig 1, S6 Table in S1 File). Additionally, both the SOFA score (AUROC: 0.747 [95% CI: 0.604-0.890]; cut-off value ≥3.5; sensitivity: 62.5%; specificity: 80.0%; P_AUROC_ = 0.003) and the CURB-65 score (AUROC: 0.776 [95% CI: 0.665-0.887]; cut-off value ≥0.5; sensitivity: 89.9%; specificity: 50.0%; P_AUROC_ = 0.001) also exhibited the good discrimination in predicting hospital mortality (Fig 1, S6 Table in S1 File). S6 Table (S1 File) summarizes the overall predictive performance of other risk factors for hospital mortality in critically ill COVID-19 patients with the Delta variant.

**Figure 1.**
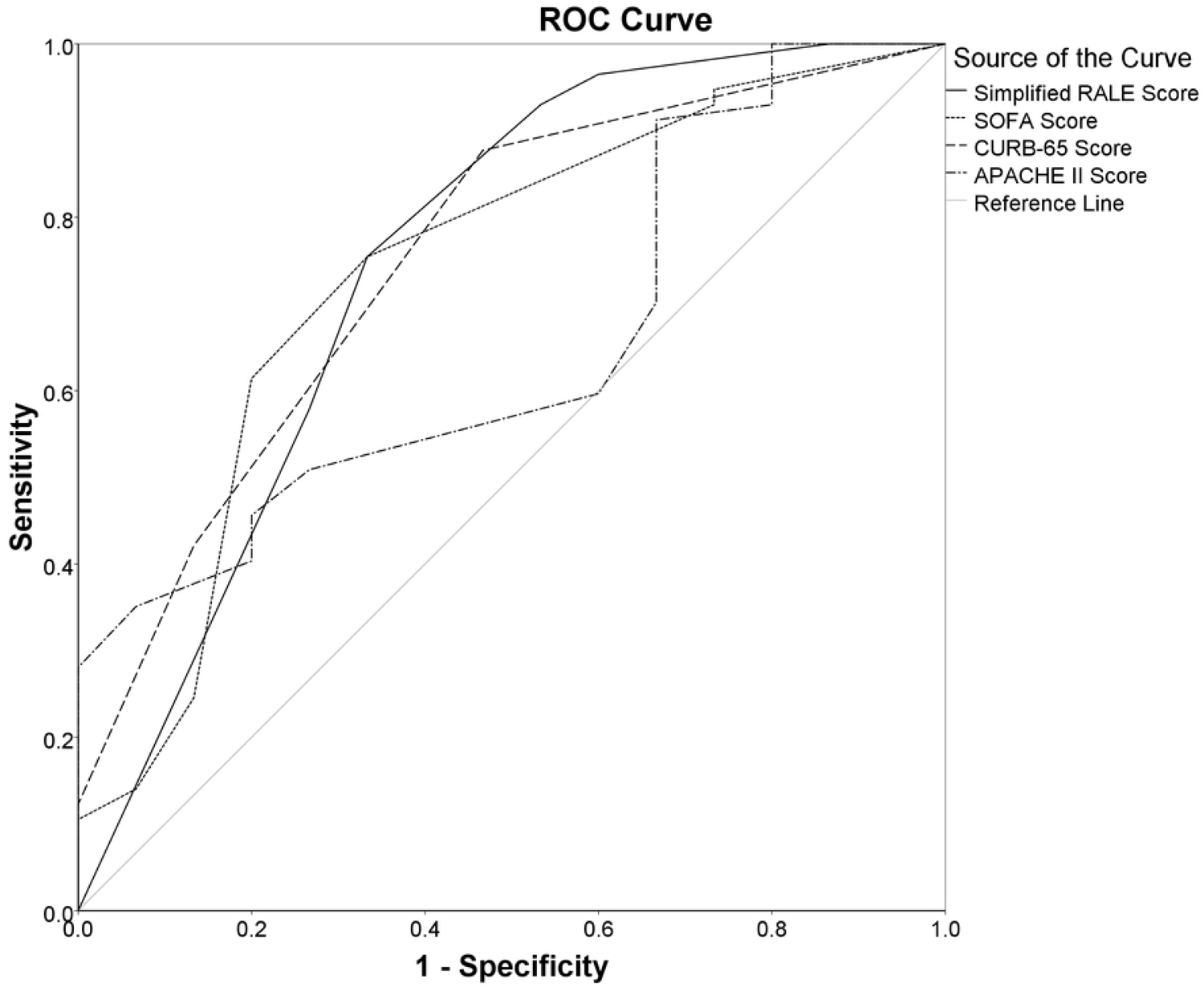
Overall predictive performance of simplified RALE, SOFA, CURB-65, and APACHE II scores for hospital mortality in critically ill COVID-19 patients with the Delta variant: The area under the ROC curves of the simplified RALE (AUROC: 0.747 [95% CI: 0.617-0.877]; cut-off value ≥5.5; sensitivity: 93.9%; specificity: 45.5%; P_AUROC_ <0.001), the SOFA score (AUROC: 0.747 [95% CI: 0.604-0.890]; cut-off value ≥3.5; sensitivity: 62.5%; specificity: 80.0%; P_AUROC_ = 0.003), CURB-65 scores (AUROC: 0.776 [95% CI: 0.665-0.887]; cut-off value ≥0.5; sensitivity: 89.9%; specificity: 50.0%; P_AUROC_ = 0.001), and APACHE II score (AUROC: 0.645 [95% CI: 0.504-0.785]; cut-off value ≥11.5; sensitivity: 33.9%; specificity: 93.8; P_AUROC_ =0.078) for predicting hospital mortality in critically ill COVID-19 patients with the Delta variant. (<u>Abbreviations</u>: **APACHE II**, Acute Physiology and Chronic Health Evaluation II; **AUROC**, areas under the receiver operating characteristic curve; **CI**, confidence interval; **CURB-65**, Confusion, Urea >7 mmol/L, Respiratory Rate ≥30 breaths/min, Blood Pressure <90 mm Hg (Systolic) or <60 mm Hg (Diastolic), Age ≥65 Years; **RALE**, Radiographic Assessment of Lung Edema; **SOFA**, Sequential Organ Failure Assessment).

S7 Table (S1 File) shows the differences between the AUROC curves among different test-pairwise, of which the AUROCs for predicting hospital mortality did not differ significantly between the simplified RALE score and SOFA score (AUROC difference: 0.015; 95% CI: - 0.117 to 0.148; Z-statistic: 0.23; p = 0.819) and the simplified RALE score and CURB-65 score (AUROC difference: 0.046; 95% CI: −0.110 to 0.203; Z-statistic: 0.58; p = 0.562). S6 Table (S1 File) also present the AUROC differences between other risk factors for hospital mortality in critically ill COVID-19 patients with the Delta variant.

### Spearman’s correlation between simplified RALE score and severity of illness

We used the Spearman’s rank correlation coefficient to examine the associations between simplified RALE score and various severity scoring systems upon admission (Table 3, S8 Table in S1 File). We observed a modest correlation between the simplified RALE score and the serum IL-6 level (Rs=0.226; p=0.032), SOFA score (Rs=0.344; p=0.002), APACHE II score (Rs=0.231; p=0.046), and CURB-65 score (Rs=0.235; p=0.019). Spearman’s correlation between other severity scoring systems upon admission is present in S8 Table (S1 File).

**Table 3.** Spearman’s correlation between the the simplified RALE Score and the severity scoring systems upon admission.

|  |  | Simplified RALE | Serum IL-6 | SOFA | APACHE II | CURB-65 | PaO<sub>2</sub>/FiO<sub>2</sub> |
| --- | --- | --- | --- | --- | --- | --- | --- |
| Simplified RALE | Rs value | 1.000 | 0.226 | 0.344 | 0.231 | 0.235 | -0.158 |
|  | P value | NA | 0.032 | 0.002 | 0.046 | 0.019 | 0.220 |
|  | N | 105 | 90 | 79 | 75 | 99 | 62 |

Abbreviations: **APACHE II**, Acute Physiology and Chronic Health Evaluation II; **CURB-65**, Confusion, Urea >7 mmol/L, Respiratory Rate ≥30 breaths/min, Blood Pressure <90 mm Hg (Systolic) or <60 mm Hg (Diastolic), and Age ≥65 Years Criteria; **IL-6**, Interleukin 6; **N**, number of patients; **NA**, not available; **PaO<sub>2</sub>/FiO<sub>2</sub>**, the ratio of partial pressure of oxygen in arterial blood to the fraction of inspiratory oxygen concentration; **Rs**, correlation coefficients; **SOFA**, Sequential Organ Failure Assessment; **RALE**, Radiographic Assessment of Lung Edema.

### Association of simplified RALE score with hospital mortality

In the univariable logistic regression analysis, the simplified RALE score ≥ 5.5 (OR: 13.000; 95% CI: 3.786-44.637; p <0.001) was significantly associated with an increased risk of hospital mortality (Table 4). In the multivariable logistic regression analysis, after accounting for confounding variables such as age (≥60 years), male gender, CCI score (≥1.5), serum IL-6 levels upon admission (≥15.8 pg/mL), SOFA score (≥3.5), and CURB-65 score (≥0.5), the simplified RALE score ≥5.5 (adjusted OR: 18.437; 95% CI: 3.215-105.741; p =0.001) remained independently associated with an increased risk of hospital mortality (Table 4). Table 4 also shows other factors that were related to hospital mortality.

**Table 4.**
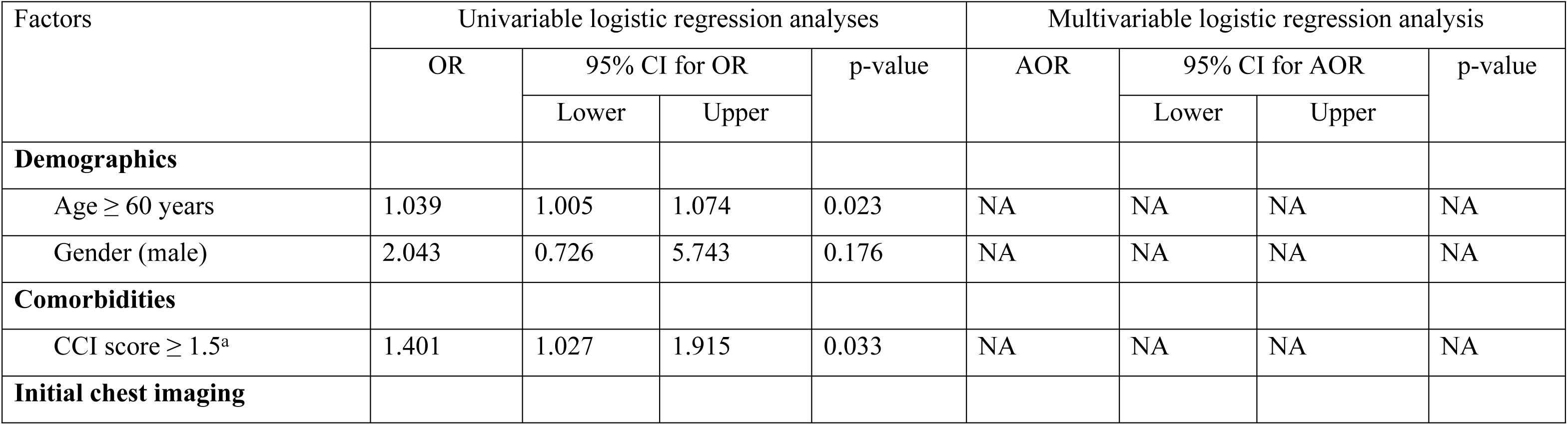
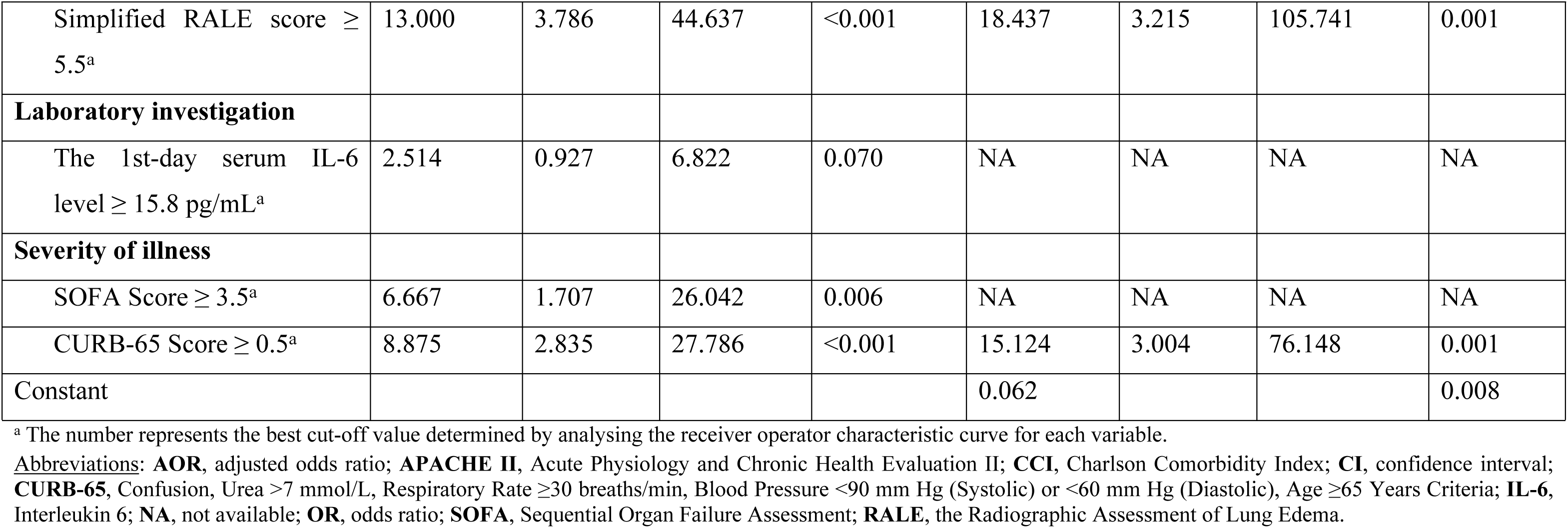
Factors associated with hospital mortality in critically ill COVID-19 patients with the Delta variant upon admission.

| Factors | Univariable logistic regression analyses |  |  |  | Multivariable logistic regression analysis |  |  |  |
| --- | --- | --- | --- | --- | --- | --- | --- | --- |
|  | OR | 95% CI for OR |  | p-value | AOR | 95% CI for AOR |  | p-value |
|  |  | Lower | Upper |  |  | Lower | Upper |  |
| **Demographics** |  |  |  |  |  |  |  |  |
| Age ≥ 60 years | 1.039 | 1.005 | 1.074 | 0.023 | NA | NA | NA | NA |
| Gender (male) | 2.043 | 0.726 | 5.743 | 0.176 | NA | NA | NA | NA |
| **Comorbidities** |  |  |  |  |  |  |  |  |
| CCI score ≥ 1.5<sup>a</sup> | 1.401 | 1.027 | 1.915 | 0.033 | NA | NA | NA | NA |
| **Initial chest imaging** |  |  |  |  |  |  |  |  |
| Simplified RALE score ≥ 5.5<sup>a</sup> | 13.000 | 3.786 | 44.637 | <0.001 | 18.437 | 3.215 | 105.741 | 0.001 |
| **Laboratory investigation** |  |  |  |  |  |  |  |  |
| The 1st-day serum IL-6 level ≥ 15.8 pg/mL<sup>a</sup> | 2.514 | 0.927 | 6.822 | 0.070 | NA | NA | NA | NA |
| **Severity of illness** |  |  |  |  |  |  |  |  |
| SOFA Score ≥ 3.5<sup>a</sup> | 6.667 | 1.707 | 26.042 | 0.006 | NA | NA | NA | NA |
| CURB-65 Score ≥ 0.5<sup>a</sup> | 8.875 | 2.835 | 27.786 | <0.001 | 15.124 | 3.004 | 76.148 | 0.001 |
| Constant |  |  |  |  | 0.062 |  |  | 0.008 |

<sup>a</sup> The number represents the best cut-off value determined by analysing the receiver operator characteristic curve for each variable.
**Abbreviations:** **AOR**, adjusted odds ratio; **APACHE II**, Acute Physiology and Chronic Health Evaluation II; **CCI**, Charlson Comorbidity Index; **CI**, confidence interval; **CURB-65**, Confusion, Urea >7 mmol/L, Respiratory Rate ≥ 30 breaths/min, Blood Pressure <90 mm Hg (Systolic) or <60 mm Hg (Diastolic), Age ≥ 65 Years Criteria; **IL-6**, Interleukin 6; **NA**, not available; **OR**, odds ratio; **SOFA**, Sequential Organ Failure Assessment; **RALE**, the Radiographic Assessment of Lung Edema.

## DISCUSSION

In the present study, we found that nearly four-fifths of critically ill COVID-19 patients with the Delta variant died in the hospital (Tables 1 and 2). Upon admission, most patients exhibited bilateral lung opacities on their CXR, with the highest occurrence of opacity distribution spanning three to four quadrants of the lungs (Table 2), a high median simplified RALE score (Table 2), and a low mean PaO_2_/FiO_2_ ratio (Table 2). In contrast to the PaO_2_/FiO_2_ ratio, serum IL-6 level, and APACHE II score (S6 Table in S1 File), the simplified RALE score exhibited a good discriminatory ability in predicting hospital mortality (Fig 1, S6 Table in S1 File). Similarly, both the SOFA score and CURB-65 score also demonstrated good discrimination in predicting hospital mortality (Fig 1, S6 Table in S1 File). Notably, there were no differences between the AUROC curves of the simplified RALE score and those of the SOFA and CURB-65 score (S7 Table in S1 File). Although the coefficients were modest, the simplified RALE score correlated with the SOFA and CURB-65 scores (Table 3). Furthermore, the present study revealed a significant association between the simplified RALE score of ≥5.5 and an increased risk of hospital mortality (Table 4). Even after adjusting for confounding factors such as age (≥60 years), male gender, CCI score (≥1.5), serum IL-6 levels upon admission (≥15.8 pg/mL), SOFA score (≥3.5), and CURB-65 score (≥0.5), the simplified RALE score of ≥5.5 remained independently associated with an increased risk of hospital mortality (Table 4).

In our study, the hospital mortality rate is higher than the rates reported in other Vietnamese studies conducted in Ho Chi Minh City (52.2%; 263/504)(15) and Binh Duong Province (64.2%; 97/151)(32), as well as an Indian study (54.6%; 306/560)(31). These disparities may arise from variations in the inclusion criteria across studies. For example, our study only included critically ill COVID-19 patients who had CXRs taken upon hospital admission. Moreover, our hospital mortality rate greatly surpasses that reported in a Swedish study (38.2%; 190/498)(33). The variation observed may stem from differences in the patients, pathogens, and clinical ability to care for critically ill patients between low- to middle-income and high-income country settings.(34–37) Additionally, the study centre almost only admitted critically ill patients with COVID-19 (92.4%, 85/92; S1 Table in S1 File) who encountered difficulties accessing treatment at lower-level hospitals, with a low rate of intubation (16.7%, 16/96; S1 Table in S1 File) and mechanical ventilation (14.0%, 14/100; S1 Table in S1 File) during transportation. Transferring critically ill patients with COVID-19 from a local to a central hospital may result in the worsening of their critical condition. Patient transfers in Vietnam may occur without intubation, ventilation, PEEP, and other medical interventions.(38–40) Although data on transfer conditions is limited, a prior survey highlighted several risk factors associated with increased patient transfers and suboptimal pre-hospital care quality.(41) Therefore, the cohort of the present study is likely to be overestimated in the hospital mortality rate.

In the early COVID-19 pandemic, the requirement to transfer a patient with COVID-19 to an ICU mainly depends on their SpO_2_, in addition to concurrent comorbidities.(6) However, there is a suggestion that this determination could also be made using imaging. Furthermore, CXRs have also been proposed as a measure for predicting the severity of COVID-19 by assessing the extent of lung involvement (7–9, 11, 12, 26), thereby offering insights into the prognosis of the infection. Although data on simple CXR severity scoring systems for COVID-19 patient mortality is scarce, a Pakistan retrospective study indicates that the initial simplified RALE score (adjusted OR: 1.278; 95% CI: 1.010–1.617) is identified as an independent predictor of hospital mortality.(11) Similarly, another Vietnamese retrospective study also shows that the simplified RALE score (OR: 1.10; 95% CI: 1.04–1.18) is significantly associated with a higher risk of hospital mortality.(12) These findings align with the present study, which highlighted the good predictive ability of the simplified RALE score in assessing hospital mortality risk (Fig 1), particularly when the score reaches ≥5.5 (Table 4). For pinpointing those most at risk of dying among critically ill COVID-19 patients, the simplified RALE score may be utilized effectively.

The present study unveiled that the PaO_2_/FiO_2_ ratio and serum IL-6 level exhibited a poor discriminatory ability in predicting hospital mortality (S6 Table in S1 File). The reliability of these measures remains a topic of ongoing debate.(42–51) A Chinese retrospective study demonstrated that the PaO_2_/FiO_2_ ratio (with an AUROC of 0.865 and a 95% CI of 0.748– 0.941) had excellent discrimination in predicting hospital mortality in COVID-19 patients requiring intensive care.(42) However, an Italian prospective study revealed a contrasting result, with the PaO_2_/FiO_2_ ratio (AUROC: 0.688, 95% CI: 0.650–0.846) showing poor discriminatory ability in predicting hospital mortality.(44) An earlier critique identified a caution problem with the PaO_2_/FiO_2_ ratio: while PaO_2_ accurately reflects a COVID-19 patient’s oxygenation, its reliability diminishes when expressed as a PaO_2_/FiO_2_ ratio.(43) As a result, inconsistent data may arise regarding the predictive value of this ratio.(42, 44) In terms of serum IL-6 level, the Chinese study found it had an excellent discriminatory ability to predict hospital mortality (AUROC: 0.900, 95% CI: 0.791–0.964).(42) However, a meta-analysis suggested that although serum IL-6 is a useful diagnostic marker for predicting severe disease, it does not appear to be associated with COVID-19 mortality.(46) Interestingly, a Belgian retrospective study observed significant differences in serum IL-6 levels between survivors and non-survivors over time, with the maximum serum IL-6 value emerging as a predictor of ICU mortality in critically ill COVID-19 patients.(47) Thus, integrating serum IL-6 into the COVID-19 prognosis warrants further exploration.

The spectrum of COVID-19 in adults ranges from asymptomatic infection to mild respiratory tract symptoms to severe pneumonia with ARDS and multi-organ dysfunction. For patients with a working diagnosis of COVID-19 pneumonia, the crucial steps in management are defining the severity of the illness and determining the most appropriate site of care. Clinicians frequently use the CURB-65 severity score due to its straightforwardness.(25) The present study found that the CURB-65 score exhibited good discrimination in predicting hospital mortality (Fig 1, S6 Table in S1 File), consistent with the finding from a comprehensive analysis of 22 predictive models applied to 411 hospitalized adults with COVID-19, which also highlighted that the CURB-65 score demonstrated good discriminatory ability in predicting 30-day mortality (AUROC: 0.75, 95% CI: 0.7–0.8)(52). Several predictive models have been suggested, but none stands out as significantly superior or precisely predicts the deterioration or mortality in critically ill patients with COVID-19.(34, 35, 52–56) Although the APACHE IV score is the most up-to-date version, some centres still use older versions, including the APACHE II score. The present study showed that the APACHE II score had poor discrimination in predicting hospital mortality in critically ill COVID-19 patients (S6 Table in S1 File), aligned with the finding of an earlier published Belgian retrospective study, which also reported a poor discriminatory ability in predicting hospital mortality using the APACHE II score (AUROC: 0.633).(56) These findings are supported by previous studies, which revealed that the APACHE II score had a good prognostic value in acutely ill or surgical patients (24, 57) but did not differentiate between sterile and infected necrotizing pancreatitis and had a poor predictive value for the severity of acute pancreatitis at 24 hours (58). However, the present study showed that the SOFA score demonstrated good discrimination in predicting hospital mortality (Fig 1, S6 Table in S1 File). In a large American cohort of patients with COVID-19 who were without mechanical ventilation within 24 hours of admission and those without a designation of do-not-resuscitate (DNR) status present at admission, the SOFA score (AUROC: 0.66, 95% CI: 0.65–0.67) exhibited a poor discriminatory ability in predicting hospital mortality.(54) This variation might be because the illness severity of our patients was worse than those of the large American cohort (mean SOFA score: 2.77 (SD: 1.91))(54). These findings are supported by an Iranian prospective study, which reported that the mean of daily SOFA scores during ICU admission (AUROC: 0.895) had excellent discrimination in predicting hospital mortality(59).

As previously mentioned, the predictive value of the test results and predictive models we discussed for assessing critically ill patients with COVID-19 remains uncertain, and the optimal utilization of these diagnostic markers and models remains unknown. However, the present study shows that the simplified RALE score, SOFA score, and CURB-65 score demonstrated good discrimination in predicting hospital mortality (Fig 1, S6 Table in S1 File), with no differences between their AUROC curves (S7 Table in S1 File). Although the coefficients were modest, the simplified RALE score correlated with the SOFA and CURB-65 scores (Table 3, S8 Table in S1 File). These findings align with previously published studies, which report that the CXR scoring system is suggested as a tool to predict the severity of COVID-19 by assessing lung involvement(7–9, 11, 12). Thus, to quantify the severity of COVID-19 pneumonia, the simplified RALE score may also be utilized effectively.

The present study has certain limitations. *Firstly*, it was conducted at a single centre in Ho Chi Minh City, Vietnam, focusing on a highly selected group of cases. The study site was urgently established in Ho Chi Minh City in response to the Delta variant epidemic, which posed significant challenges for medical providers dealing with a large influx of COVID-19 patients. Additionally, all COVID-19 patients were transferred from various hospitals within the Ho Chi Minh City area, and the treatments they received before admission influenced their disease severity upon enrolment. As a result, these elements hinder the seamless integration of pre-hospital and hospital treatment procedures, as well as the collection of clinical data for surveillance, quality enhancement, and research purposes. Additionally, they introduce an implicit selection bias and incomplete patient inclusion in the study database, which could potentially result in an overestimation of the mortality rate. *Secondly*, we conducted the present study during the Delta variant epidemic in Vietnam; however, no included patients with COVID-19 underwent genomic analysis to confirm infection with a SARS-CoV-2 mutant virus strain. However, according to sequencing data at the time of the study period in Ho Chi Minh City, COVID-19 was almost entirely caused by one lineage of the Delta variant (AY.57).(60) In that sense, we evaluated a relatively homogeneous patient population in virologically. *Finally*, although the sample size was large enough, the confidence interval was wide (±8.02%), which might influence the normal distribution of the collected sample. Further studies with larger sample sizes might be needed to consolidate the conclusions.

## CONCLUSIONS

This study investigated a highly selected cohort of critically ill COVID-19 patients with the Delta variant, a high simplified RALE score, and a high mortality rate presented to an Intensive Care Centre for the Treatment of Critically Ill Patients with COVID-19 in Ho Chi Minh City, Vietnam. Beyond its ability to quantify severity and predict hospital mortality, the simplified RALE score also emerged as an independent predictor of hospital mortality. For pinpointing those most at risk of progressing and dying among critically ill COVID-19 patients, the simplified RALE score can be utilized effectively.

## Data Availability

All relevant data are within the manuscript and its Supporting Information files.

## Acknowledgements

We are grateful to the special task teams from the Bach Mai Hospital and Vietnam’s Ministry of Health and to all the healthcare heroes across the country who helped Ho Chi Minh City during the third wave of the COVID-19 pandemic in Vietnam. We also appreciate the support and statistical advice from the staff of the Faculty of Public Health at the Thai Binh University of Medicine and Pharmacy. Finally, we thank Miss Truc-Cam Nguyen from Stanford University, Stanford, California, the United States of America, and Miss Mai Phuong Nguyen from the Hotchkiss School, Lakeville, Connecticut, the United States of America, for their support of our manuscript.

## Supporting information

S1 File. Supplementary Results

